# Covid-19 Will Reduce US Life Expectancy at Birth by More Than One Year in 2020

**DOI:** 10.1101/2020.12.03.20243717

**Authors:** Patrick Heuveline

## Abstract

On December 3^rd^, 2020, the cumulative number of U.S. Covid-19 deaths tallied by Johns Hopkins University (JHU) online dashboard reached 275,000, surpassing the number at which life table calculations show Covid-19 mortality will lower the U.S. life expectancy at birth (LEB) for 2020 by one full year. Such an impact on the U.S. LEB is unprecedented since the end of World War II. With additional deaths by the year end, the reduction in 2020 LEB induced by Covid-19 deaths will inexorably exceed one year. Factoring the expected continuation of secular gains against other causes of mortality, the U.S. LEB should still drop by more than a full year between 2019 and 2020. By comparison, the opioid-overdose crisis led to a decline in U.S. LEB averaging .1 year annually, from 78.9 years in 2014 to 78.6 years in 2017. At its peak, the HIV epidemic reduced the U.S. LEB by .3 year in a single year, from 75.8 years in 1992 to 75.5 years in 1993. As of now, the US LEB is expected to fall back to the level it first reached in 2010. In other words, the impact of Covid-19 on U.S. mortality can be expected to cancel a decade of gains against all other causes of mortality combined.

---

On December 3^rd^, 2020, the cumulative number of U.S. Covid-19 deaths tallied by Johns Hopkins University (JHU) online dashboard^1^ reached 275,000. As a proportion of the U.S. population, this number amounts to 83 Covid-19 deaths per 100,000 persons. A more salient measure of the mortality impact of the pandemic may be that this cumulative number now exceeds 273,500 deaths, a threshold at which life table calculations show Covid-19 mortality will lower the U.S. life expectancy at birth (LEB) for 2020 by one full year.

The most recent estimate of the U.S. LEB is 78.7 years for the year 2018, an increase of one-tenth of a year over the previous annual estimate.^2^ This small increase is fairly typical of contemporary annual increases in LEB, in the order of .2 years on average in the U.S. and other high-income nations,^3^ that secular gains against the leading causes of mortality provide in these nations. Severe public health crises have occasionally caused reversals in this upward trend in LEB. The most recent example is the opioid-overdose crisis in the U.S., which led to a decline in LEB averaging .1 year annually, from 78.9 years in 2014 to 78.6 years in 2017. At its peak, the HIV epidemic reduced the U.S. LEB by .3 year in a single year, from 75.8 years in 1992 to 75.5 years in 1993.^4^

The impact of Covid-19 on the U.S. LEB is unprecedented, however, since the end of World War II. With additional deaths by the year end, the reduction in 2020 LEB induced by Covid-19 deaths will inexorably exceed one year. As shown on Figure 1, the current year-end projections from the University of Washington’s Institute for Health Metrics and Evaluation (IHME)^5^ translate into a reduction of 1.20 to 1.25 years. Factoring the expected continuation of secular gains against other causes of mortality, the U.S. LEB should still drop by more than a full year between 2019 and 2020. As of now, the US LEB is actually expected to fall back to 78.7 years,^6^ a level it first reached in 2010. In other words, the impact of Covid-19 on U.S. mortality can be expected to cancel a decade of gains against all other causes of mortality combined.

**Figure 1:**
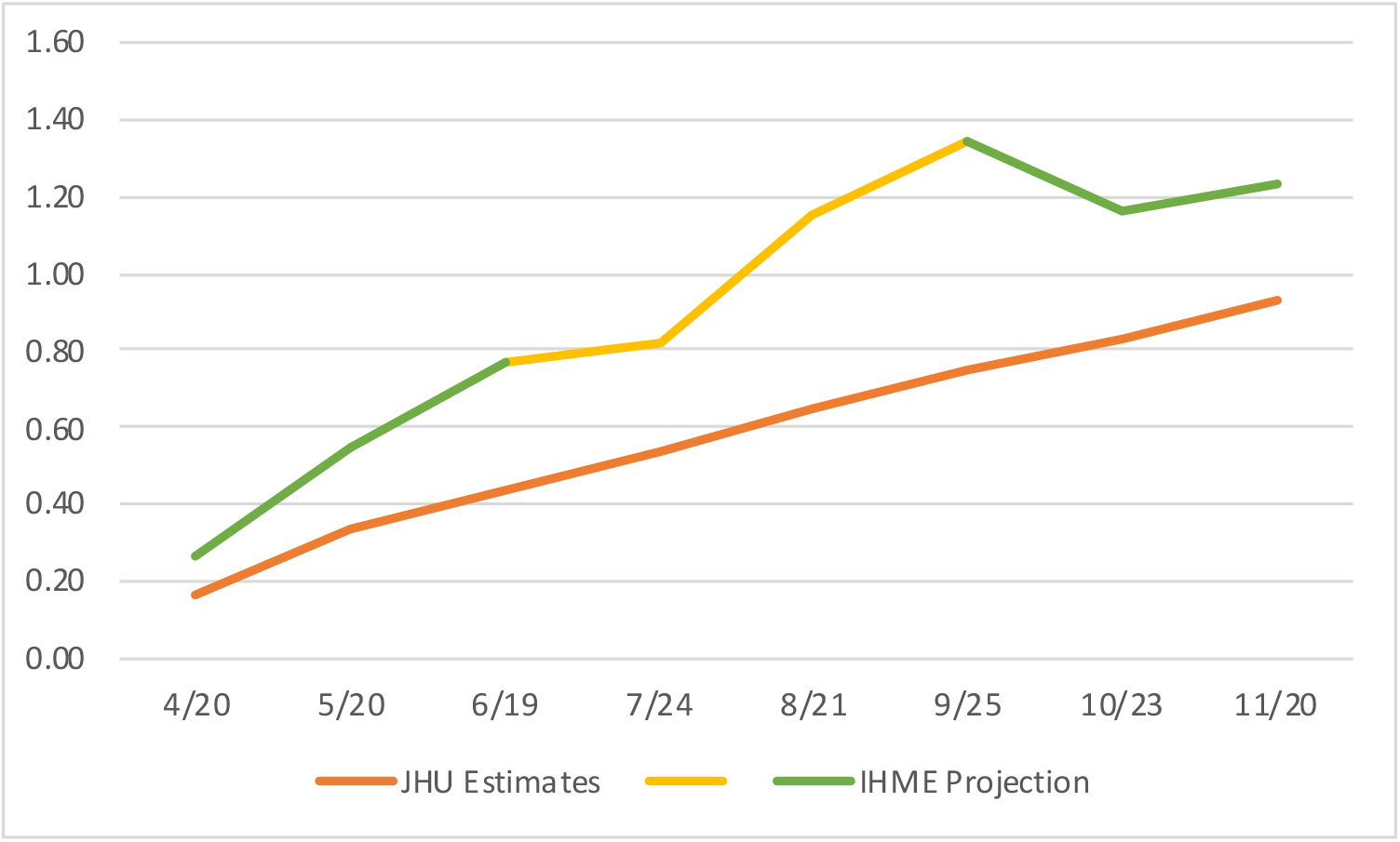
Impact of Covid-19 Deaths on the U.S. LEB in 2020, Based on Year-To-Date Estimates (JHU) and Projections (IHME)^a^ over Time. Sources. Impact based on JHU estimates: author’s calculations. Impact based on IHME projections: Heuveline and Tzen, Github repository.^7^ _a_ The increase in the projected impact from late June to late September (yellow trend line above) is due in part to gradual extensions of the projection period. Up to June, projections are for deaths until August 4^th^, 2020. From September on, projections are for deaths until January 1^st^, 2021.

## Discussion

Due to the lag between trends in Covid-19 cases and deaths, the uncertainty concerning the number of confirmed Covid-19 deaths by year end is now rather modest. A greater source of uncertainty concerns the extent to which Covid-19 deaths to date have been fully accounted for. The difference between the actual number of deaths and the number expected had past conditions continued to prevail, the number of excess deaths in the US is about 25% higher than the estimated number of Covid-19 deaths.^8^ Some might be deaths from Covid-19 for which the cause of death was not properly identified. Others might be deaths from other causes that indirectly resulted from conditions Covid-19 contributed to (e.g., saturation of local hospital capacity). Either way, these excess deaths represent the true impact of Covid-19 on mortality and LEB.

The threshold number of 273,500 is also an estimate derived under standard life table assumptions. Among them, the assumption that mortality risks from different causes are independent is unlikely to hold entirely.^9^ As individuals dying of Covid-19 might have been more likely to die of other causes than those who survived the risk of Covid-19 mortality, the impact of Covid-19 on LEB should be less than estimated under the assumption of independence and the threshold number of deaths be higher than 273,500. Several underlying conditions are known to increase the risk of death from Covid-19, however, the remaining life expectancy for an average individual dying from Covid-19 and for an average individual of the same age and sex do not appear to be drastically different. One study in the UK, for instance, estimated that the average number of years of life lost was reduced from 13 years per Covid-19 death to 12 years (average for both sex) when controlling for several comorbidities known to increase the risk of Covid-19 mortality.^10^ The fact that the ratio of excess deaths to confirmed Covid-19 deaths has remained relatively stable over time also counters the notion that most individuals who succumbed to Covid-19 were already very frail and likely to have died of other causes within a few weeks or months. Overall, the evidence to date overwhelmingly suggests that if both the threshold number of 273,500 deaths and the December-3^rd^ tally of 275,000 deaths are underestimates, the latter is underestimated to a greater extent than the threshold number. It thus appears highly likely that the threshold number of deaths required to reduce the U.S. LEB by a one full year in 2020 has indeed been reached.

## Methods

The reduction in 2020 LEB induced by Covid-19 deaths is assessed by comparing, for each sex, two life tables representing the mortality conditions expected to prevail in 2020 with and without Covid-19 mortality. The first one can be derived by interpolation from the United Nations projections of life table values for the 2015-20 and 2020-25 periods.^11^ To derive the second one from the first one is to reverse the demographic technique use to calculate an associated decrement-deleted life table from a multiple-decrement life tables. This involves calculating new age-specific survival probabilities for which Chiang^12^ provided an elegant solution:

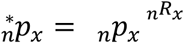

where _*n*_*p*_*x*_ and _*n*_^*\**^*p*_*x*_ are the survival probabilities in the two life tables and _*n*_*R*_*x*_ is the ratio of projected deaths in 2020 with and without Covid-19 deaths. New values of _*n*_*a*_*x*_, the age-specific number of years lived after age *x* for individuals dying in the age interval *x* to *x*+*n*, also need to be derived, for which Preston et al.^13^ suggest the interpolation:

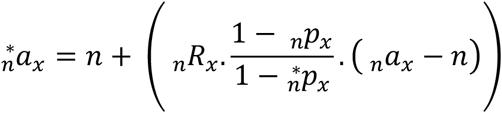

## Data Availability

All data are from available online sources

https://coronavirus.jhu.edu/map.html

https://covid19.healthdata.org/united-states-of-america?view=total-deaths&tab=trend

## Acknowledgment

The author benefited from facilities and resources provided by the California Center for Population Research at UCLA (CCPR), which receives core support (P2C-HD041022) from the Eunice Kennedy Shriver National Institute of Child Health and Human Development (NICHD).

## References

1. Dong E, Du H & Gardner L. An interactive web-based dashboard to track COVID-19 in real time. The Lancet Inf Dis 20(5): 533. https://www.thelancet.com/journals/laninf/article/PIIS1473-3099(20)30120-1/fulltext

2. Xu JQ, Murphy SL, Kochanek KD & Arias E. 2020. Mortality in the United States, 2018. NCHS Data Brief 355(January 2020). Hyattsville, MD: National Center for Vital Statistics. https://www.cdc.gov/nchs/data/databriefs/db355-h.pdf

3. Ho JY & Hendi AS. 2018. Recent Trends in Life Expectancy across High Income Countries: Retrospective Observational Study. BMJ 362: 2562. https://www.bmj.com/content/362/bmj.k2562

4. Arias E & Xu JQ. 2019. United States life tables, 2017. NVSR 2019, 68(7). Hyattsville, MD: National Center for Vital Statistics. https://www.cdc.gov/nchs/data/nvsr/nvsr68/nvsr68_07-508.pdf

5. University of Washington, Institute for Health Metrics and Evaluation. COVID-19 projections. https://covid19.healthdata.org/. Last used November 28,2020.

6. Heuveline P & Tzen M. 2020. Beyond Deaths per Capita: Comparative CoViD-19 Mortality Indicators. MedRxiv First published online May 5,2000. https://www.medrxiv.org/content/10.1101/2020.04.29.20085506v8. Last used July 14, 2020

7. https://github.com/statsccpr/ind-cov-mort

8. Weinberger, DW, Chen J, Cohen, T et al. 2020. Estimation of Excess Deaths Associated With the COVID-19 Pandemic in the United States, March to May 2020. JAMA Intern Med. Published online July 1, 2020. doi:10.1001/jamainternmed.2020.3391

9. Heuveline P. 2020. The Mean Unfulfilled Lifespan (MUL): A New Indicator of a Disease Impact on the Individual Lifespan. MedRxiv First published online September 17, 2000. https://www.medrxiv.org/content/10.1101/2020.08.09.20171264v3

10. Hanlon P et al. 2020. COVID-19 – Exploring the Implications of Long-Term Condition Type and Extent of Multimorbidity on Years of Life Lost: A Modelling Study. Welcome Open Res 5: 75. https://doi.org/10.12688/wellcomeopenres.15849.1

11. United Nations, Department of Economic and Social Affairs, Population Division. World Population Prospects 2019, Online Edition. Rev. https://population.un.org/wpp/Download/Standard/Mortality/. Last used June 2, 2020.

12. Chiang, C. L. 1968. An Introduction to Stochastic Processes in Biostatistics. New York: Wiley.

13. Preston SH, Heuveline P & Guillot M. Demography: measuring and modeling population processes. Blackwell: Malden, MA, Oxford, England & Carlton, Australia, 2001.

